# Clinical manifestation and laboratory parameters associated with progression to severe dengue in children: a systematic review and meta-analysis

**DOI:** 10.1101/2023.01.24.23284985

**Authors:** Indra Sandinirwan, Bani Muslim, Henry Leo, Hasanah Hasanah, Permata Putri Karina

## Abstract

**Background:** The ingenuity to predict the progression to severe dengue is crucial in managing dengue patients. The previous meta-analysis has been performed on adults, and none has been performed specifically on children. We conducted a systematic review and meta-analysis to determine the clinical manifestations and laboratory parameters associated with the progression to severe dengue according to WHO criteria.

**Methods:** We focused on searching six medical databases for studies published from Jan 1, 2000, to Dec 31, 2020. The meta-analysis used random-effects or fixed-effects models to estimate pooled effect sizes. We also assessed the heterogeneity and publication bias. This study was registered with PROSPERO, CRD42021224439.

**Results:** We included 49 of papers in the systematic review, and we encased the final selected 39 papers comprising 23 potential predictors in the meta-analyses. Among 23 factors studied, seven clinical manifestations demonstrated association with disease progression in children, including neurological signs, gastrointestinal bleeding, clinical fluid accumulation, hepatomegaly, vomiting, abdominal pain, and petechiae. Six laboratory parameters are associated during the early days of illness, including elevated hematocrit, elevated aspartate aminotransferase [AST], elevated alanine aminotransferase [ALT], low platelet count, low albumin levels, and elevated activated partial thromboplastin time. Dengue virus serotype 2 (DENV-2) and secondary infections were also associated with severe disease progression.

**Conclusion:** This finding supports the use of the warning signs described in the WHO 2009 guidelines. In addition, monitoring serum albumin, AST/ALT levels, identifying infecting dengue serotypes, and immunological status could improve the risk prediction of disease progression.

## Introduction

Dengue is still a public health concern across the globe. The global incidence is estimated to be 390 million individuals infected each year, with mortality rates ranging from 10,000 to 20,000 per year.^1,2^ Dengue infection can affect all ages, including adults and children. Dengue mortality will increase as the disease progress to severe dengue conditions or dengue shock syndrome, especially in children with comorbid factors. Although most patients have mild symptoms, a small proportion will progress to severe dengue or dengue hemorrhagic fever, which can be life-threatening. This progression to the severe disease commonly occurs between ddays4 and 6 of illness.^3^

During hospitalization, clinical and laboratory monitoring for these patients should always be carried out. Serial hematocrit and platelet examinations have become the standard examinations in various health care centers. To assist clinicians in the early detection of severe disease progression, World Health Organization (WHO) has issued guidelines for dengue, namely the 1997 and 2009 WHO Dengue Guidelines.^3,4^ Clinical symptoms such as persistent vomiting, abdominal pain, bleeding manifestations, and fluid accumulation are warning signs associated with the occurrence of severe dengue. Several laboratory parameters associated with dengue disease severity include hematocrit, platelets, aspartate aminotransferase, alanine aminotransferase, lactate dehydrogenase, and serum albumin.^5^

This systematic review aimed to identify whether specific clinical manifestations and laboratory parameters have a strong, moderate, or low association with severe dengue. In addition, the authors hoped that the results of this study could help early decision-making to optimize patient management and improve the quality of care for children.

## Methods

### Protocol and Registration

This systematic review was conducted based on the Preferred Reporting Items for Systematic Review and Meta-Analysis (PRISMA) Statement^6^ and a guide to systematic review and meta-analysis of prognostic factor studies.^7^ The protocol of this systematic review has been registered in The International Prospective Register of Systematic Reviews (PROSPERO) database (CRD42021224439).

### Eligibility Criteria

We aimed to include observational studies of pediatric dengue patients who were clinically and serologically confirmed and were treated inpatient or outpatient in a hospital, health center, or teaching hospital. The dengue criteria used in this study were according to the 1997 or 2009 WHO dengue classification. Studies should show an association between clinical manifestations and laboratory parameters with severe dengue prevalence and the minimal study size of at least twenty patients. To be included in systematic reviews and meta-analyses, papers must provide a summary measure or effect measure for severe dengue in the form of odds ratio (OR), risk ratio (RR), with p-values or confidence intervals (CI) or have to give crude data that allowed calculation of a measure. Severe dengue in our study was a confirmed dengue patient who was in a clinical shock state due to plasma leakage, severe bleeding, or severe organ involvement. Published articles in the last 20 years (2000 to 2020) were expected to provide novelty and updates on existing systematic reviews and meta-analyses. Only papers written in the English language were included. We excluded prognostic factors related to gene expression, cell receptors, virological studies, neutralizing antibodies, cytokines & plasma proteins. We eliminated conference abstracts/papers, supplementary issues, review articles, seroepidemiological studies, adult population studies, and brief reports.

### Information sources, search strategy, and study selection

From December 2020 to February 2021, we conducted an article search from six electronic databases. The search strategy combines “dengue fever” and “children” with possible synonyms. The search terms and the study selection process are shown in Table 1 and Figure 1. We also searched for related studies through reference and citation checks. Bramer et al. suggest that to get optimal results in a systematic review, the author at least does a search on Embase, MEDLINE/PubMed, Web of Science, and Google Scholar to ensure adequate coverage.^8^ During the article search process, we found a small number of articles whose full text was not in English, we decided not to include them. We also did not include papers from gray literature and unpublished studies that have not gone through the peer-review process. We contacted the corresponding author by email regarding information that needs to be confirmed in the article obtained.

**Table 1.** Search queries of the systematic review

| Databases | Search Query | Hits |
| --- | --- | --- |
| PubMed | Dengue OR Severe dengue OR Complicated dengue OR Predictor OR Predictive OR Prognostic AND Pediatric NOT Adult Filters: Observational Study, Child: birth-18 years | 1170 |
| Embase | Dengue AND Children | 102 |
| Google Scholar | Dengue Prospective Cohort Children OR Pediatric OR Paediatric OR Pediatrics OR Paediatrics<br>-Adult -Zika -Pneumonia -Vaccine -Economic -Chikungunya -Malaria -Sepsis | 583 |
| Web of Science | Dengue AND Children OR Pediatric OR Paediatric | 7 |
| EBSCO | Dengue fever OR dengue syndrome AND children OR adolescents OR youth OR child OR teenager OR teens OR young people OR kids OR paediatric OR pediatric | 82 |
| Scopus | Dengue AND Predictors AND Children | 130 |

**Figure 1.**
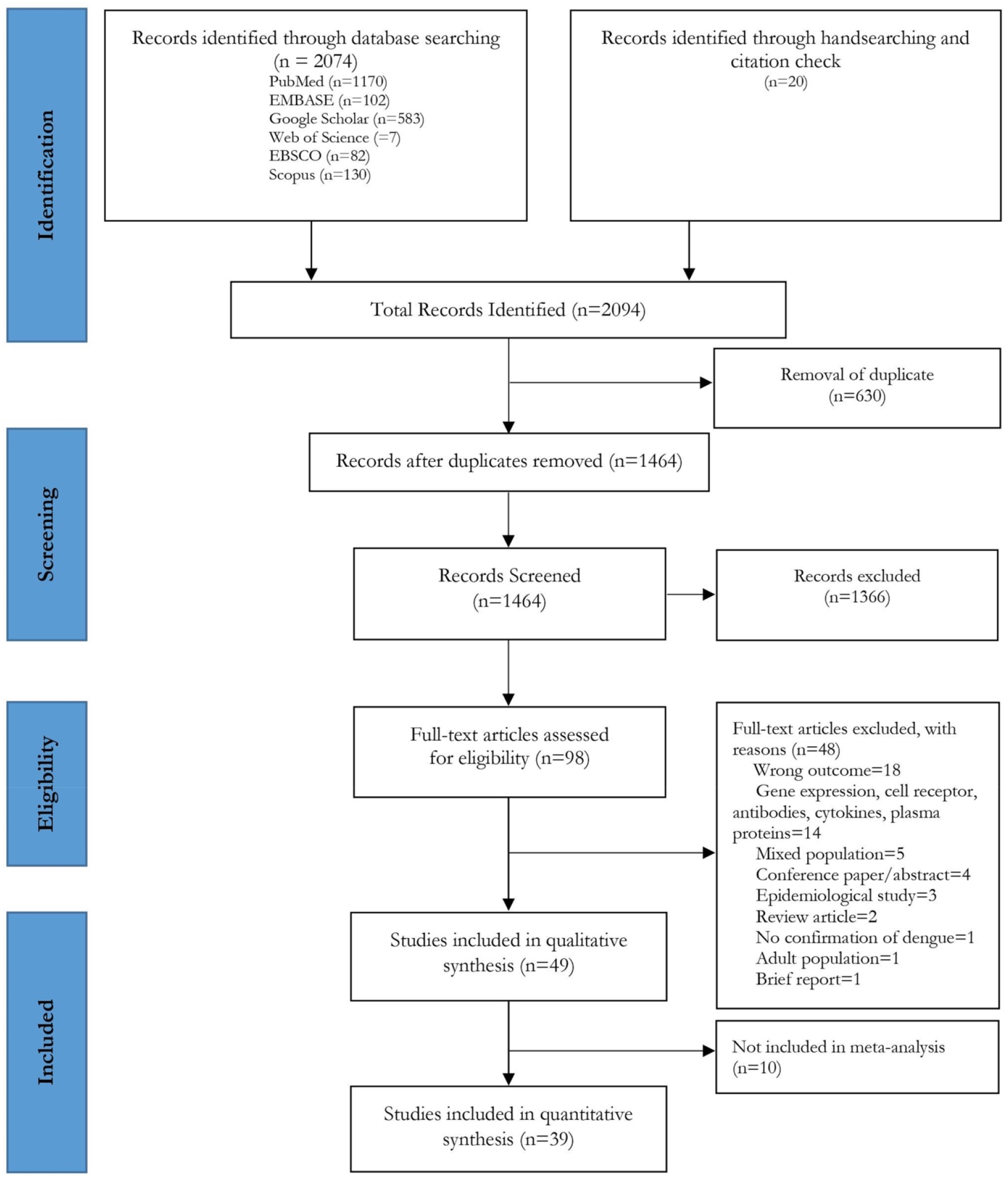
Flowchart for article selection

We used Zotero software to remove the article duplicates and exported the listed articles to the Rayyan app for systematic review.^9^ The titles and abstracts were independently reviewed by two authors (IS and BM) based on pre-determined eligibility criteria. After screening and removing irrelevant articles, we managed to list 98 articles. The same two authors conducted an overall text assessment after retrieving full-text papers from each publisher. Finally, the third reviewer (HL) resolved the differences between the two reviewers, and the final decision was determined by consensus.

### Data items and data collection process

The primary information presented was the association between clinical manifestations and specific laboratory parameters with the severity of dengue infection. In addition, we extracted information about the health facilities and the place where the research was conducted, the time of the study, the name of the first author, the year of publication, study design and methods, study size, and population characteristics in each study. Data from the included studies were extracted using a standardized, pre-piloted form. The data from eligible studies were entered into Microsoft Excel, and the two authors independently carried out the extraction process (IS, BM). Disagreements were resolved by a third author (HL).

### Risk of bias in individual studies

Two authors independently (IS and HL) examined the risk of bias using the Quality in Prognostic Studies (QUIPS) tools.^10^ Disagreements resolved by a third author (HH). The risk of bias assessment results was presented using Microsoft Excel.

### Synthesis of results

All included studies were presented in a narrative and summary table. For each prognostic factor (clinical manifestation or laboratory parameter) evaluated in at least two studies, we summarized it in a forest plot and submitted it to the meta-analysis. Meta-analysis was performed using Comprehensive Meta-Analysis software version 3.0 (Biostat, USA) and MetaXL version 5.3 (EpiGear, Australia). Combining the classification of dengue based on the 1997 and 2009 WHO guidelines, we divided the progression of dengue into two: patients with severe disease (dengue hemorrhagic fever, dengue shock syndrome according to 1997 WHO guidelines or severe dengue according to WHO 2009 guidelines) and patients without the severe disease forms (dengue fever, non-severe dengue, and dengue with warning sign).

The pooled odds ratio indicates the strength of the association between certain prognostic factors and severe dengue. Dichotomous variables and continuous variables were analyzed to calculate the effect size (in the form of odds ratio or standardized mean difference) if there were DSS/DHF and DF groups (WHO 1997 criteria) or severe dengue and non-severe dengue/dengue with warning sign groups (WHO 2009 criteria). The SMD value was then converted into the OR based on the Borenstein method. The two variables (both dichotomous and continuous) were combined to obtain a pooled OR to increase the number of studies included in the analysis.^11–13^

Current guidelines for prognostic studies recommend reporting both crude and adjusted association measures.^14^ The adjusted effect sizes can be obtained if a study uses a multivariate analysis approach. However, most of the eligible studies in our systematic review used univariate analysis, and only a small proportion used multivariate analysis. We decided to use only the crude measures of association in this meta-analysis. We performed a meta-analysis with fixed effects and random effects for the different predictors to generate pooled estimates. To further investigate the robustness of pooled estimates, sensitivity analysis was also performed by removing studies with extreme effect size and heterogeneity. We also did a subgroup analysis categorized by the study’s country

The heterogeneity assessment was conducted using the Cochran Q statistic and *I*^2^ statistic. Studies were considered heterogeneous if the p-value for the Cochran Q was less than 0.1; the *I*^2^ expressed the proportion of variation across studies that is due to heterogeneity. The level of heterogeneity was categorized as low (0-25%); low to moderate (25% to <50%); moderate to high (50% to <75%); and high (≥75%).^15,16^ The agreement between reviewers in study selection was assessed with Cohen’s kappa.

To detect any publication bias, we used Funnel plots and Doi plots. The symmetrical Doi Plot indicates the absence of publication bias, while the asymmetric plot indicates the presence of publication bias, considering the Luis Furuya-Kanamori (LFK) index. In the funnel plot, if Egger’s test shows p<0.05, it indicates publication bias.^17^ LFK index up to ±1 indicates no asymmetry, values that exceed ±1 but did not exceed ±2 indicate minor asymmetry, and if the value exceeds ±2 shows major asymmetry. Unlike the Funnel plots requiring a minimum of 10 studies, Doi plots can be used for fewer studies.^16,18^ To enhance the symmetry, we also used Duval and Tweedie’s trim and fill method to assess the sensitivity of the crude estimates to publication bias.^19^ Factors that were only investigated in one study or for which raw data could not be combined, the association of these factors with disease progression was derived from the original research and presented narratively.

## Results

### Characteristics of the selected studies

Of the 2094 papers identified, 49 were included in the systematic review ^20–68^ (Figure 1), with the agreement between the two reviewers at 93% (Cohen’s kappa = 0.72). All included studies are observational studies [100%, (49/49)] and among the studies, [78%, (38/49)] were prospective cohort, [14%, (7/49)] were retrospective cohort, [4%, (2/49)] were case-control, and [4%, (2/49)] were cross-sectional study. Most of the included studies were conducted in Asia and Latin America, published between 2000 to 2020. The countries included in this study are India, Thailand, Indonesia, Vietnam, Sri Lanka, Paraguay, Nicaragua, Brazil, and the Philippines. Only one study took populations from seven dengue-endemic countries, namely a study conducted by Rosenberger et al.^36^ Almost all of the studies comprised inpatient (hospitalized) populations, and only one study conducted by Tuan et al.^57^ incorporated outpatient study populations.

The inclusion criteria for patients varied between studies but generally included pediatric patients with fever symptoms for 1–3 days (≤72 hours) as inclusions. About 57% (28 out of 49) of the studies defined severity using “1997 WHO classification”, while the remaining studies [21% (21/49)] used “2009 WHO classification”. The supplementary information shows the details and summary of included studies (S1 Table and S2 Table).

The total number of patients in the 49 studies included in this systematic review was 93628 patients, with a pooled prevalence of severe dengue was 0.30 (95%CI 0.25–0.34) (S1 Text). The management of dengue in pediatric patients follows the WHO guidelines, or at a more specific level, adjusted to the health service policy at the hospital where the study was conducted.

### Risk of bias

We used the QUIPS tool to examine the risk of bias in the six domains (Figure 2). Most studies have a low risk of bias in study participation, study attrition, prognostic factor measurement, outcome measurement, and statistical analysis and reporting. In the confounding study domain, we found a moderate risk of bias. Details of the risk of bias assessment are presented in the supplementary information (S3 Table).

**Figure 2.**
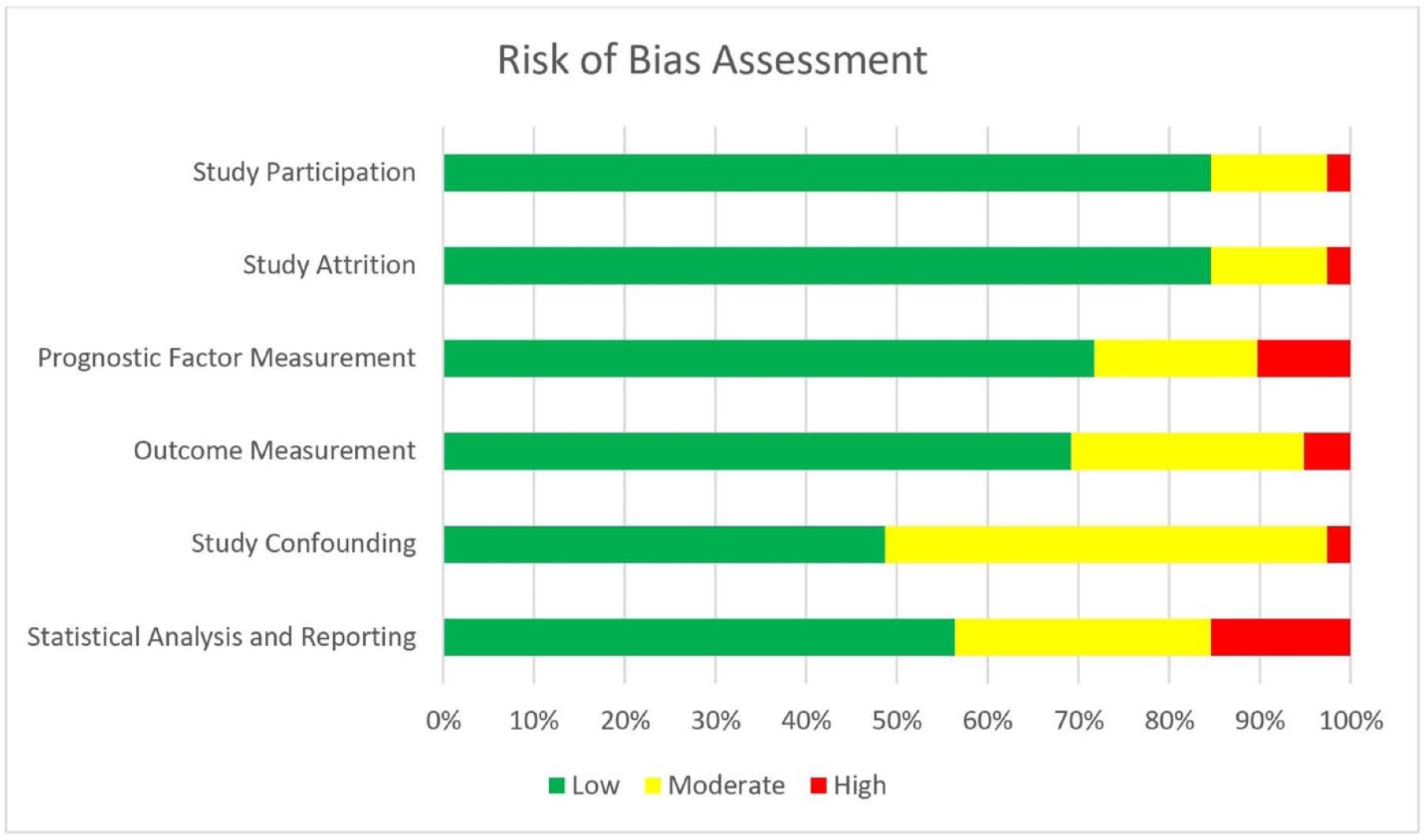
Risk of bias assessment according to the six domains of the Quality in Prognostic Studies (QUIPS) tool for 39 observational studies included in the meta-analysis

### The prognostic factor of severe dengue

Twenty-three potential prognostic factors (reported in 39 studies) were evaluated in at least three studies and were submitted to the meta-analysis. An overview of prognostic factors evaluated is presented in Table 2, and the summary of crude and adjusted ORs of significant prognostic factors can be found in Table 3. In addition, the individual forest plots, Doi plots, Funnel plots, sensitivity analyses, and subgroup analyses for each prognostic factor are given in supplementary information (S2 Text).

**Table 2.** Overview of prognostic factor evaluated

| No | Factor | Number of Studies | Number Significant | Model | Association with Severe Dengue |  | Heterogeneity |  | Egger's 2-tailed bias p-value | LFK index | Adjusted OR (95% CI) |
| --- | --- | --- | --- | --- | --- | --- | --- | --- | --- | --- | --- |
|  |  |  |  |  | Pooled OR (95% CI) | *p-value* | I<sup>2</sup> | *p-value* |  |  |  |
| 1 | Normal nutrition | 2 | 0 | Fixed | 1.02 (0.60 – 1.75) | 0.930 | 0 | 0.402 | - | - | - |
| 2 | Male gender | 16 | 4 | Random | 1.04 (0.88 – 1.24) | 0.641 | 60 | 0.001 | 0.02 | 4.40 | <sup>a</sup> 0.89 (0.57 – 1.05)<br><sup>b</sup> 1.21 (1.05 – 1.39) I<sup>2</sup>=0% |
| 3 | Age | 18 | 6 | Random | 1.00 (0.83 – 1.20) | 0.844 | 91 | <0.001 | 0.86 | 1.23 | <sup>b</sup> 0.97 (0.81 – 1.17) I<sup>2</sup>=91% |
| 4 | Neurological Sign | 9 | 4 | Random | 6.33 (2.34 – 17.13) | <0.001 | 68 | 0.002 | 0.04 | 1.76 | <sup>a</sup> 4.11 (1.59 – 10.65)<br><sup>b</sup> 6.88 (2.91 – 16.25) I<sup>2</sup>=36% |
| 5 | Gastrointestinal bleeding | 7 | 2 | Random | 5.34 (1.67 – 17.01) | 0.005 | 46 | 0.083 | 0.06 | -3.30 | <sup>b</sup> 5.87 (2.03 – 16.98) I<sup>2</sup>=0% |
| 6 | Clinical fluid accumulation | 18 | 15 | Random | 4.66 (2.08 – 10.45) | <0.001 | 95 | <0.001 | 0.04 | 2.43 | <sup>a</sup> 3.86 (1.77 – 8.40)<br><sup>b</sup> 3.03 (1.28 – 7.17) I<sup>2</sup>=95% |
| 7 | Hepatomegaly | 19 | 13 | Random | 2.64 (1.76 – 3.94) | <0.001 | 88 | <0.001 | 0.008 | 3.78 | <sup>a</sup> 2.09 (1.41 – 3.10)<br><sup>b</sup> 2.28 (1.54 – 3.38) I<sup>2</sup>=88% |
| 8 | Vomiting | 14 | 8 | Random | 2.01 (1.54 – 2.64) | <0.001 | 48 | 0.021 | 0.52 | 0.47 | - |
| 9 | Abdominal Pain | 15 | 8 | Random | 1.91 (1.26 – 2.88) | 0.002 | 82 | <0.001 | 0.23 | 3.48 | <sup>b</sup> 1.58 (1.07 – 2.35) I<sup>2</sup>=81% |
| 10 | Petechiae | 7 | 4 | Random | 1.79 (1.29 – 2.49) | <0.001 | 44 | 0.097 | 0.14 | 4.09 | <sup>b</sup> 1.62 (1.31 – 2.02) I<sup>2</sup>=12% |
| 11 | Positive Tourniquet Test | 8 | 4 | Random | 0.97 (0.46 – 2.05) | 0.306 | 89 | <0.001 | 0.74 | 0.37 | - |
| 12 | Rash | 5 | 2 | Fixed | 1.09 (0.82 – 1.46) | 0.536 | 74 | 0.004 | 0.13 | -3.26 | <sup>b</sup> 0.71 (0.47 – 1.06) I<sup>2</sup>=55% |
| 13 | Elevated Hematocrit | 20 | 11 | Random | 1.76 (1.50 – 2.07) | <0.001 | 89 | <0.001 | <0.001 | 7.04 | <sup>a</sup> 1.17 (0.99 – 1.39)<br><sup>b</sup> 3.14 (2.03 – 4.85), I<sup>2</sup>=81% |
| 14 | Low Platelet Count | 23 | 13 | Random | 2.01 (1.70 – 2.38) | <0.001 | 90 | <0.001 | <0.001 | 8.94 | <sup>a</sup> 1.48 (1.25 – 1.75)<br><sup>b</sup> 1.76 (1.50 – 2.06) I<sup>2</sup>=89% |
| 15 | Higher WBC | 11 | 4 | Random | 1.01 (0.76 – 1.35) | 0.940 | 66 | 0.001 | 0.68 | -0.54 | - |
| 16 | Low Albumin Levels | 11 | 7 | Random | 6.03 (2.34 – 15.53) | <0.001 | 96 | <0.001 | 0.002 | 8.51 | <sup>b</sup> 7.34 (3.29 – 16.38) I<sup>2</sup>=88% |
| 17 | Elevated AST | 15 | 10 | Random | 3.06 (2.00 – 4.69) | <0.001 | 91 | <0.001 | 0.03 | 3.70 | <sup>a</sup> 3.63 (2.33 – 5.67)<br><sup>b</sup> 3.08 (2.18 – 4.36) I<sup>2</sup>=68% |
| 18 | Elevated ALT | 11 | 5 | Random | 2.68 (1.59 – 4.51) | 0.001 | 81 | <0.001 | 0.21 | 2.14 | <sup>b</sup> 1.98 (1.27 – 3.08) $I^2=73\%$ |
| 19 | Temperature | 8 | 3 | Random | 0.73 (0.43 – 1.22) | 0.231 | 81 | <0.001 | 0.012 | -5.2 | <sup>a</sup> 1.08 (0.65 – 1.81)<br><sup>b</sup> 0.58 (0.39 – 0.88) $I^2=0\%$ |
| 20 | DENV-2 Serotype | 4 | 1 | Fixed | 1.66 (1.30 – 2.13) | <0.001 | 1 | 0.387 | 0.89 | 0.15 | - |
| 21 | Activated Partial Thromboplastin Time | 4 | 3 | Fixed | 8.74 (4.79 – 15.95) | <0.001 | 83 | 0.001 | 0.85 | 1.13 | <sup>b</sup> 4.59 (2.24 – 9.37) $I^2=70\%$ |
| 22 | Secondary Infection | 5 | 4 | Fixed | 2.02 (1.64 – 2.49) | <0.001 | 89 | <0.001 | 0.14 | 4.56 | <sup>a</sup> 1.59 (1.31 – 1.93)<br><sup>b</sup> 8.66 (4.99 – 15.04) $I^2=56\%$ |
| 23 | Primary Infection | 4 | 2 | Fixed | 0.52 (0.41 – 0.67) | <0.001 | 83 | <0.001 | 0.54 | -2.56 | <sup>b</sup> 0.62 (0.48 – 0.81) $I^2=0\%$ |

- a) Adjusted odds ratio by enhancing the symmetry using the trim and fill method of Duval and Tweedie b) Adjusted odds ratio by sensitivity analysis

**Table 3.** Summary of crude and adjusted odds ratio of significant prognostic factors submitted to meta-analysis

| Prognostic factor | Number of Studies | Pooled OR (95%CI) | I<sup>2</sup> (%) | LFK index |
| --- | --- | --- | --- | --- |
| Neurological sign |  |  |  |  |
| All studies | 9 | 6.33 (2.34 – 17.13) | 68 | 1.76 (minor asymmetry) |
| Sensitivity analysis | 7 | 6.88 (2.91 – 16.25) | 36 | 0.07 (no asymmetry) |
| Gastrointestinal bleeding |  |  |  |  |
| All studies | 7 | 5.34 (1.67 – 17.01) | 46 | -3.30 (major asymmetry) |
| Sensitivity analysis | 5 | 5.87 (2.03 – 16.98) | 0 | 0.58 (no asymmetry) |
| Clinical fluid accumulation |  |  |  |  |
| All studies | 18 | 4.66 (2.08 – 10.45) | 95 | 2.43 (major asymmetry) |
| Sensitivity analysis | 14 | 3.03 (1.28 – 7.17) | 95 | 0.87 (no asymmetry) |
| Hepatomegaly |  |  |  |  |
| All studies | 19 | 2.64 (1.76 – 3.94) | 88 | 3.78 (major asymmetry) |
| Sensitivity analysis | 16 | 2.28 (1.54 – 3.38) | 88 | *2.49 (major asymmetry) |
| Vomiting |  |  |  |  |
| All studies | 14 | 2.01 (1.54 – 2.64) | 49 | 0.47 (no asymmetry) |
| Abdominal pain |  |  |  |  |
| All studies | 15 | 1.91 (1.26 – 2.88) | 82 | 3.48 (major asymmetry) |
| Sensitivity analysis | 12 | 1.58 (1.07 – 2.35) | 81 | *1.60 (minor asymmetry) |
| Petechiae |  |  |  |  |
| All studies | 7 | 1.79 (1.29 – 2.49) | 44 | 4.09 (major asymmetry) |
| Sensitivity analysis | 5 | 1.62 (1.31 – 2.02) | 12 | 0.07 (no asymmetry) |
| Elevated hematocrit |  |  |  |  |
| All studies | 20 | 1.76 (1.50 – 2.07) | 89 | 7.04 (major asymmetry) |
| Sensitivity analysis | 16 | 3.14 (2.03 – 4.85) | 81 | *2.63 (major asymmetry) |
| Low platelet count |  |  |  |  |
| All studies | 23 | 2.01 (1.70 – 2.38) | 90 | 8.94 (major asymmetry) |
| Sensitivity analysis | 20 | 1.76 (1.50 – 2.06) | 89 | *7.77 (major asymmetry) |
| Low albumin levels |  |  |  |  |
| All studies | 11 | 6.03 (2.34 – 15.53) | 96 | 8.51 (major asymmetry) |
| Sensitivity analysis | 10 | 7.34 (3.29 – 16.38) | 88 | -0.27 (no asymmetry) |
| Elevated aspartate aminotransferase |  |  |  |  |
| All studies | 15 | 3.06 (2.00 – 4.69) | 91 | 3.70 (major asymmetry) |
| Sensitivity analysis | 10 | 3.08 (2.18 – 4.36) | 68 | 0.00 (no asymmetry) |
| Elevated alanine aminotransferase |  |  |  |  |
| All studies | 11 | 2.68 (1.59 – 4.51) | 81 | 2.14 (major asymmetry) |
| Sensitivity analysis | 9 | 1.98 (1.27 – 3.08) | 73 | 0.24 (no asymmetry) |
| Elevated activated partial thromboplastin time |  |  |  |  |
| All studies | 4 | 8.74 (4.79 – 15.95) | 83 | 1.13 (minor asymmetry) |
| Sensitivity analysis | 3 | 4.59 (2.24 – 9.37) | 70 | 0.68 (no asymmetry) |
| DENV-2 serotype |  |  |  |  |
| All studies | 4 | 1.66 (1.30 – 2.13) | 1 | 0.15 (no asymmetry) |
| Secondary infection |  |  |  |  |
| All studies | 5 | 2.02 (1.64 – 2.49) | 89 | 4.56 (major asymmetry) |
| Sensitivity analysis | 3 | 8.66 (4.99 – 15.04) | 56 | *-1.75 (minor asymmetry) |

\*No further LFK index approaching zero beyond this point to eliminate the asymmetry

**Table 4.** Summary of factors associated with progression to severe dengue

| Association with progression | Factors |
| --- | --- |
| Very Strong | Neurological signs, gastrointestinal bleeding, clinical fluid accumulation (ascites or pleural effusion), low albumin levels, and elevated activated partial thromboplastin time. |
| Moderate to Strong | Hepatomegaly, elevated aspartate aminotransferase, elevated alanine aminotransferase. |
| Weak to Moderate | Vomiting, abdominal pain, petechiae, low platelet count, elevated hematocrit, secondary infection, DENV-2 serotype. |
| No Association | Normal nutritional status, male gender, age, temperature, positive tourniquet test, white blood cells, viremia. |

The age profile of pediatric patients with severe dengue was no different from those without severe dengue. Heterogeneity in pooled results is very high (91%), and after removing studies with extreme effect sizes in sensitivity analysis, the heterogeneity remains unchanged. The OR value for the age factor was 1.00 (95%CI 0.83 – 1.20), and no evidence of publication bias (Egger’s test p=0.86). The association between gender and progression to severe disease was not significant. Moderate to high heterogeneity (*I*^2^=60%) and publication bias was found in studies examining the male gender as a predictor of severe dengue (Egger’s test p=0.02). The result of the pooled OR was 1.04 (95%CI 0.88 – 1.24).

Three studies reported an association between nutritional status and progression to severe dengue. The study conducted by Baiduri et al. showed a strong relationship between overweight-obesity and progression to severe dengue with an RR of 94 (95%CI 4.47 – 1989) through a multivariate analysis approach. While the study conducted by Tantracheewathorn et al. and Dewi et al., normal nutritional status did not show an association with progression to severe dengue. After including two studies in the meta-analysis, we obtained an OR of 1.02 (95%CI 0.60 – 1.75) with no heterogeneity (*I*^2^=0%).

In terms of clinical manifestations, we found that neurologic sign factors are strongly associated with the progression of severe dengue. Neurological manifestations may include drowsiness, convulsions, decreased consciousness, or lethargy. The meta-analysis of nine studies yielded a high pooled OR of 6.33 (95%CI 2.34 – 17.13) with moderate to high heterogeneity (*I*^2^=68%) and publication bias (Egger’s test p=0,04).

Bleeding manifestation can be mild or severe. Almost all studies include petechiae as a predictive factor for disease progression and gastrointestinal bleeding, either as melena or hematemesisins its extreme form. Children having petechiae will have an increased risk of developing the severe disease with an OR value of 1.79 (95%CI 1.23 – 2.49), with low to moderate heterogeneity (*I*^2^=44%), no evidence of publication bias (Egger’s test p=0.14) but with major asymmetry (LFK=4.09). For children who presented with gastrointestinal bleeding, the risk of progression to severe dengue was increased with an OR of 5.34 (95%CI 1.67 – 17.01), low to moderate heterogeneity (*I*^2^=46%), without evidence of publication bias (Egger’s test p=0,06).

For clinical fluid accumulation, whether pleural effusion or ascites, 18 studies were included in the meta-analysis and gave an OR value of 4.66 (95%CI 2.08 – 10.45), with evidence of high heterogeneity (*I*^2^=95%) and publication bias (Egger’s test p=0.04). The positive relationship between clinical fluid accumulation and progression to severe disease was also consistent in sensitivity analysis with an OR of 3.03 (95%CI 1.28 – 7.17). Thus, the evidence of high heterogeneity in this factor remains, but no asymmetry was found (LFK index 0.87).

Hepatomegaly was moderately associated with severe dengue after pooling 19 eligible studies. High heterogeneity was found (*I*^2^=88%) as well as publication bias (Egger’s test p=0.008), with an OR of 2.64 (95%CI 1.76 – 3.94). We found that vomiting was associated with an increased risk of progression to severe dengue. The definition of vomiting is not clearly defined in most studies. A significant association between vomiting and disease progression was given with an OR of 2.01 (95%CI 1.54 – 2.63) with low to moderate heterogeneity (*I*^2^=48%) and no evidence of publication bias (Egger’s test p=0.52). Abdominal pain was also associated with progression to severe disease with an OR of 1.91 (95%CI 1.26 – 2.88), with evidence of high heterogeneity (*I*^2^=82%) and no publication bias was found (Egger’s test p=0.23).

Without evidence of publication bias, rash and positive tourniquet tests were not associated with progression to severe dengue. Heterogeneity was high in the positive tourniquet test and was moderate to high in the pooled results of positive rash signs. An elevated body temperature was not associated with progression to severe dengue with evidence of high heterogeneity (*I*^2^=81%) and publication bias (Egger’s test p=0.012), given an OR of 1.73 (95%CI 0.43 – 1.22).

Hematological parameters, including hematocrit, platelet count, and white blood cell count,t were included in the meta-analysis. Of those, hematocrit and platelet count were significantly associated with progression to severe dengue, while white blood cells were not. By pooling 23 studies related to low platelet count, we obtained an OR of 2.01 (95% CI 1.70 – 2.38) with evidence of high heterogeneity (*I*^2^=90%) and publication bias (Egger’s test p<0.001).

Elevated hematocrit was identified as a positive association with severe dengue. By pooling 20 studies related to hematocrit, we obtained an OR of 1.76 (95%CI 1.50 – 2.07) with evidence of high heterogeneity (*I*^2^=89%) as well as publication bias (Egger’s Test p<0.001).

Eleven studies were included in the meta-analysis of serum albumin. All studies have consistently reported that children with lower albumin levels have an increased risk of developing severe dengue. Low albumin levels gave an OR of 6.03 (95%CI 2.34 – 15.53) with evidence of high heterogeneity (*I*^2^=96%) and publication bias (Egger’s test p=0.002). One study that was not submitted in the meta-analysis, conducted by Kularatnam et al.,^31^ showed that the reduction in serum albumin levels seen on the third and fourth days of illness were valid predictors of entering into the critical phase in dengue infection.

Fifteen studies were included in the meta-analysis related to aspartate aminotransferase and eleven studies related to alanine aminotransferase. This meta-analysis assessing the association between AST and ALT levels with disease progression showed that higher of these enzymes were associated with progression to severe disease. Evidence of heterogeneity in the two factors was high, with publication bias on the AST factor (Egger’s test p=0.003) but not on the ALT factor (Egger’s test p=0.21). The pooled OR for the AST factor was 3.06 (95% CI 2.00 – 4.69), and the pooled OR for the ALT factor was 2.68 (95%CI 1.59 – 4.51) with evidence of moderate to high heterogeneity (*I*^2^=73%) without asymmetry (LFK index 0.24).

The association between activated partial thromboplastin time (aPTT) factor with disease progression showed a strong positive association. By including four studies, the pooled OR was 8.74 (95% CI 4.79 – 15.95) with evidence of high heterogeneity (*I*^2^=83%) but without publication bias (Egger’s test 0.85). One study not submitted in the meta-analysis by Budastra et al.^60^ found that aPTT values may be used as a predictor for bleeding manifestation in dengue hemorrhagic fever using a cox regression approach with an RR of 2.02 (95%CI 1.92 – 3.90, p=0.02).

Secondary infection with the dengue virus was significantly associated with progression to severe dengue. In five studies, the pooled OR was 2.02 (95% CI 1.64 – 2.49) with evidence of high heterogeneity (*I*^2^=89) but no publication bias (Egger’s test 0.14). Primary infection with dengue virus was inversely associated with progression to severe dengue in the meta-analysis. The pooled OR of this factor was 0.52 (95%CI 0.41 – 0.67) with high heterogeneity (*I*^2^=83%) and no publication bias (Egger’s test p=0.54). A study conducted by Endy et al.^45^ showed that cross-reactive memory humoral immune responses appear beneficial in symptomatic secondary DENV-3 infection but not in secondary DENV-2 or DENV-1 infection. Poeranto et al.^67^ showed that only DENV-3 could cause severe clinical manifestations in primary infection. In addition, a secondary infection caused by DENV-1, which is considered to induce mild symptoms, involves the risk of severe manifestation, especially if DENV-2 was the cause of the primary infection.

Included studies in the meta-analysis have shown that the DENV-2 serotype was associated with severe disease progression. Pooling results from four studies gave an OR of 1.66 (95%CI 1.30 – 2.13) with evidence of low heterogeneity and no publication bias (Egger’s test p=0.54). Consistent findings were also reported by Lovera et al.,^32^ whose study showed that the DENV serotype profoundly impacts the clinical manifestations and the dengue severity. In this article, the DENV-2 infections were more frequently associated with the requirement of fluid resuscitation, shock, and more prolonged hospital stay.

A study conducted by Sirikutt et al.^44^ showed that serum lactate and lactate dehydrogenase (LDH) was found to be elevated in DHF and/or DSS patients. Lactate might be used as a predictor of DSS if the level was >2 U/L on day 0. Moreover, LDH can be used to predict severe dengue or DSS if the level propagated to approximately 1000 IU on day 0 of illness. A study by Yacoub et al.^28^ has shown that initial venous lactates in dengue patients on the first day of admission to ICU were associated with severe outcomes of recurrent shock and respiratory distress. Lactate levels correlated with the total intravenous fluids received but did not correlate with other hemodynamic parameters.

A study conducted by Chaiyaratana et al.^49^ showed that a serum ferritin level ≥1200 ng/mL has a high sensitivity but a relatively low specificity to predict the occurrence of severe dengue. Bongsebandhu et al.^50^ showed that D-dimer was significantly associated with dengue severity. Early increasing D-dimer in the febrile stage could predict the severity of dengue infection during the initial phase of the illness with a 68.4% positive predictive value. Suvarna et al.^37^ found that lipid profile changes accompany dengue infection, some of which may indicate severity.

Regarding viremia, the study conducted by Tuan et al.^57^ through a multivariate logistic model found that viremia magnitude was independently associated with severe dengue. Still, the viremia itself was not included in the final prognostic model of Early Severe Dengue Identifier (ESDI). The four factors included in the ESDI model were vomiting, platelet count, AST levels, and NS1 rapid test status. The study conducted by Singla et al.^56^ showed that dengue viremia was not associated with disease severity. Dengue viremia was indistinguishable between patients with non-severe dengue or severe dengue either in primary or secondary infection. However, all the secondary infections with the recurrence bleeding had a significantly higher viremia than primary infections despite showing clinical improvement from severe dengue. Van Ta et al.^64^ showed that DENV concentration/viremia within the first day of hospitalization (≤72 hours) could not be used as a prognostic factor of DSS. The DENV concentration was highest on day-2 of fever and higher in the secondary infection group, consistent with other studies.

## Discussion

In this systematic review and meta-analysis, we found that age, gender, and normal nutritional status were not associated with the development of severe dengue. Meanwhile, neurological signs, gastrointestinal bleeding, clinical fluid accumulation, hepatomegaly, vomiting, abdominal pain, and petechiae were associated with severe dengue. In addition, low platelet count, elevated hematocrit, low albumin levels, elevated AST/ALT, elevated aPTT, DENV-2 serotype, and secondary infection were also associated with progression to severe disease.

Age was not associated with progression to severe dengue in our study, similar to the findings reported by Zhang et al.^69^ In another meta-analysis,^70^ the factor of young age in children was associated with disease severity, and the interpretation may be more convincing because of its dose-response meta-analysis.

Most studies did not categorize nutritional status as malnourished, normal nutrition, overweight, or obese. Several studies determine body weight as a variable and its value on a continuous scale. To more accurately determine the nutritional status of children, it is necessary to consider both their weight and height or even other indicators such as upper arm circumference or skinfold thickness.^71^ In other words, the association between nutritional status with progression to severe dengue from this meta-analysis should be interpreted cautiously. The meta-analysis conducted by Sangkaew et al.^70^ in the adult population showed that nutritional status was not associated with severe dengue. Meanwhile, a study conducted by Trang et al.,^72^ showed that children with normal nutrition were inversely associated with DSS compared to DHF (OR: 0.87, 95 % CI: 0.77–0.99). Our review has not been able to find a sufficient number of articles related to obesity and dengue severity. A study conducted by Zulkipli et al. ^73^ seems to have conclusive findings that obesity is a risk factor for severe dengue in children.

In most studies, some clinical symptoms that have a strong association include neurological signs, which are represented by symptoms of drowsiness, convulsion, alteration in sensorium, or lethargy. Gastrointestinal bleeding, described in the form of melena or hematemesis, is also strongly associated with severe forms of the disease. Milder forms of bleeding such as petechiae that appear early during illness are also prognostic signs of disease progression. Clinical fluid accumulation manifesting as ascites or pleural effusion are two clinical manifestations that can be particularly important considering the hallmark of plasma leakage in dengue pathophysiology. Thus, this prognostic sign will guide clinicians promptly for further patient management.

We also found that clinical manifestations of vomiting and abdominal pain were associated with disease progression. These two subjective symptoms that are frequently complained about by patients were also reported to be significant in other studies.^5,69,70^ The criteria for vomiting in most studies are not described clearly. However, according to the study conducted by Vuong et al.,^74^ the criteria for persistent vomiting should be two times vomiting or more per day.

Numerous patients with severe dengue exhibited manifestation of severe plasma leakage only. In contrast, a small proportion of patients who develop severe bleeding or severe organ dysfunction, which occurred less frequently, often exhibited a higher degree of overlap with severe plasma leakage. In addition, patients with severe bleeding without leakage may be predisposed to bleeding disorders.^36^ Hence, this potential confounder should be considered because the predisposition to bleeding disorders has not been explored further in the included studies.

The association between secondary infection and disease progression is well documented and represents an antibody-dependent enhancement mechanism. A modeling study conducted by Clapham et al.^75^ showed that severe manifestations were found in 40% (95% CI 0.36–0.45) of secondary infections and i8% (95% CI 0.16–0.20) of primary infections. DENV-2 serotype was significantly associated with disease progression in children, and these findings were consistent with previous meta-analyses.^5,70^

Hepatic manifestations in dengue patients result from direct viral toxicity or dysregulated immunologic injury. The spectrum of hepatic involvement can be mild or even severe transaminases elevation in the form of acute liver failure.^76^ Our analysis showed that high AST or ALT levels in the early stages of illness were significantly associated with severe dengue. The studies included in our meta-analysis have varying cut-off values of AST/ALT concentrations, which might cause the high heterogeneity. However, based on most of the average AST and ALT reported in the included studies, we suggested that concentrations higher than three times the upper limit of normal value were associated with severe disease progression.

In our study, albumin levels in the first days of illness were significantly lower in patients with severe dengue than those with non-severe dengue. During plasma leakage, albumin was also extravasated. However, patients’ nutritional status influenced their initial albumin levels, and albumin levels were generally lower in undernourished children.^77^ In most of the included studies, low albumin levels were described as less than 3.5 g/dL. Our meta-analysis is in line with the recommendations of the 2011 SEARO dengue guidelines, in which patients whose albumin levels are <3.5 g/dL or undergoing a decrease of 0.5 g/dL from baseline during the febrile phase are associated with disease progression.^78^

This meta-analysis has some limitations. As we searched for studies reported in the English language, some of the studies published in non-English languages fortunately also have their English versions, making it possible for data extraction. However, some studies might be excluded due to language limitations. In addition, we did not search studies from gray literature. Both events may contribute to publication bias. The predisposing genetic factors were also not included in the meta-analysis because these have not been routinely analyzed along with the clinical outcomes. Analysis between clinical outcomes and predisposing genetic factors is quite challenging. Nevertheless, a study has evaluated multiple genetic variants that confer clinically prominent risk for disease progression, namely a study conducted by Pare et al.^79^ More than one-third of the prognostic factors have high heterogeneity while others were moderate to high. However, in sensitivity analysis and subgroup analysis, the effect sizes are quite similar and give consistent results with evidence of lowered heterogeneity.

In our study, we found that neurological signs, gastrointestinal bleeding, clinical fluid accumulation, low albumin levels, elevated aPTT, hepatomegaly, elevated AST/ALT, vomiting, abdominal pain, petechiae, low platelet count, elevated hematocrit, secondary infection, and DENV-2 serotypes were associated with disease progression. This finding supports the use of the warning signs (abdominal pain, persistent vomiting, clinical evidence of fluid accumulation, bleeding, lethargy and/or restlessness, liver enlargement, rise in hematocrit with rapid decrease in platelet count) described in the WHO 2009 guidelines. In addition, monitoring serum albumin, AST/ALT levels, identifying infecting dengue serotypes, and immunological status could improve the risk prediction of disease progression further.

## Data Availability

All data produced in the present study are available upon reasonable request to the authors

## Supporting information

***S1 Table***. *Details of the observational studies included. (DOCX)*

***S2 Table***. *Summary of the studies included. (DOCX)*

***S3 Table***. *Risk of bias assessment for 49 studies using the QUIPS tool. (XLSX)*

***S1 Text***. *The pooled prevalence of severe dengue in children. (DOCX)*

***S2 Text***. *Forest plot, Doi plot, Funnel plot, sensitivity analysis, and subgroup analysis (DOCX)*

***S3 Text***. *Preferred Reporting Items for Systematic Review and Meta-Analyses (PRISMA) Checklist (DOCX)*

## Author Contribution

Idea and study design: IS; Data collection and analysis: IS, BM, HL; Article draft writing: IS, BM, HH; Draft revision: BM, PPK; Writing supervision: PPK. All authors have read the manuscript and have approved this submission.

## Acknowledgments

Thanks to all of the participants in this article.

